# The scope and needs related to training, research, and mentorship among early-career addiction medicine professionals: an online global survey followed by expert group discussions

**DOI:** 10.1101/2022.03.24.22272872

**Authors:** Roshan Bhad, Sophia Achab, Parnian Rafei, Preethy Kathiresan, Hossein Mohaddes Ardabili, Jenna L. Butner, Laura Orsolini, Katrine Melby, Mehdi Farokhnia, Venkata Lakshmi Narasimha, Kelly Ridley, Serenella Tolomeo, Mitika Kanabar, Beatrice Matanje, Paolo Grandinetti, ISAM NExT Consortium, Marc N. Potenza, Hamed Ekhtiari, Alexander Baldacchino

## Abstract

Addiction medicine is a developing field, with many young professionals opting for a career in this area. However, globally, early-career professionals often face challenges in this field, such as lack of competency-based training due to a shortage of trainers, low availability of institutions with appropriate infrastructure, and limited resources for adequate training, particularly in developing countries. On the other hand, in developed countries, early career professionals may struggle with mentorship, limited job opportunities, and challenges with establishing a suitable research area.

The International Society of Addiction Medicine (ISAM) New Professionals Exploration, Training & Education (NExT) committee, a global platform for early-career addiction medicine professionals (ECAMPs), conducted an online survey using a modified Delphi-based approach among ECAMPs across 56 countries to assess and understand the need and scope for standardized training, research opportunities, and mentorship. The survey was conducted in 2 phases. A total of 110 respondents participated in Phase I (online key informant survey), and 28 respondents participated in Phase II (online expert group discussions on the three themes identified in Phase I). Most participants agreed with the lack of standardized training, structured mentorship programmes, research funding, and research opportunities in addiction medicine for ECAMPs. There is a need for standardized training programmes, improving research opportunities, and effective mentorship programmes to promote the next generation of addiction medicine professionals and further development to the entire field. The efforts of ISAM-NExT are well-received and give a template of how this gap can be addressed.

## 1. Introduction

Addiction medicine is a relatively new field of medicine, with a growing number of early career professionals (ECPs) opting for a career in this area. However, there are several challenges in terms of lack of well-structured training shortage of institutes with infrastructure for adequate training and trainers, i.e., formally trained mental health and medical professionals with experience in addiction medicine (1, 2). These limitations prevent ECPs in several countries from pursuing a career in addiction medicine. There are limited resources and training opportunities for ECPs in upper-middle (UMICs) and lower-middle-income countries (LMICs). In high-income countries (HICs), where there is no or less dearth of experts and infrastructure, the challenges include receiving appropriate mentorship and choosing a suitable research area (3–5). Therefore, there is a need for a global platform helping early-career addiction medicine professionals (ECAMPs), including trainees, and connecting them with each other and with trainers and mentors worldwide. There is also a need to facilitate the launch and implementation of standardized training programmes, creating research and education opportunities, as well as fellowships and mentorship programmes in each subspecialty of the addiction medicine field (6). These needs become extremely necessary mainly due to the significant variability in the standards and quality of training programmes in the field of addiction and/or psychiatry globally, which is a major challenge for many ECAMPs in many countries (7).

Similarly, the assessment of training in addiction medicine and/or psychiatry is limited. For example, the International Certification in Addiction Medicine by the International Society of Addiction Medicine (ISAM) is one of the few examples of a well-established association able to provide global standards in validating and certifying knowledge in addiction medicine for professionals. However, currently, the examination by ISAM includes assessment for theory (based on multiple-choice questions (MCQs), without any practical exam or real case vignettes (8, 9). Hence, it is essential first to identify and clearly understand the needs and the demand for a standardized assessment of training in addiction medicine and/or psychiatry to develop and implement a universal curriculum in addiction training programmes.

Building research collaboration across the globe and developing a practice-based research network is of high importance, given the eclectic nature of the field of addiction medicine and its significance is emphasized by policymakers and various global organizations, as well. Unfortunately, there are hardly any international organizations and networks in addiction medicine that address the need for researchers on a global platform for research collaboration. The National Institute on Drug Abuse (NIDA) is an organization that provides research support for early and mid-career addiction professionals; however, support is often limited to United States-based researchers and institutes (10). Assessing the need and scope for research opportunities exclusively for ECAMPs will inform policymakers regarding various issues and challenges. Quality mentoring and strategic planning, along with a favorable environment, are some of the elements that should be combined to create a successful career in research (11). Moreover, there is a need to assess deficiencies in training, research interest, and need for mentoring among early-career addiction professionals and address important issues that may help them in career development to mid-career. This may motivate and encourage ECAMPs to take up addiction medicine as an informed career choice since they can see the career trajectory and growth prospects ahead. The ISAM NExT (New Professionals Exploration, Training & Education) committee was established in 2020 with the primary objective of increasing and improving the capacity of addiction medicine training and other educational activities among ECAMPs. The committee constitutes 30 early career addiction professional members, including a chair and two co-chairs from 22 countries. We conducted a two-phase global cross-sectional online survey among ECAMPs to understand the need and scope for standardized training, research opportunities, and mentorship in the field of addiction medicine.

## 2. Methods

### 2.1. Study design

A two-phase global online survey was conducted using a mixed-method, modified Delphi-based approach (12, 13). The first phase of the survey was carried out using an online survey in which 270 ECAMPs were approached. The first round of the survey took place from October 2020 to March 2021, and the results were finalized in April 2021. The second phase of the study comprised three focus group discussions to obtain consensus on key themes elicited in the first phase.

An online Google survey tool was prepared by the research team for phase I (available as supplementary material). Eligible participants (ECAMPs) were identified (sample of convenience) across different regions of the world using membership directory of professional societies in the field and social media/research networks, i.e., ResearchGate and LinkedIn enrolled. ECAMPs (n=270) were then invited to participate in the study via email. Total of 125 respondents provided written informed consent for participation in study on email in the first phase. Written informed consent were also obtained from participants prior to enrollment in the second phase of the study. to Subsequently, the data were analyzed, and the recommendations were compiled based on feedback from a core group of 13 collaborators of the ISAM NExT expert committee for the research project.

### 2.2. Inclusion and exclusion criteria

For the purpose of this study, the operational definition of “Early Career Addiction Medicine Professionals (ECAMPs)” has been used (Table 1) (14) who were clinicians, scholars, resident doctors, and professionals working in or with an interest in the field of addiction medicine within 10 years of obtaining MD/MSc/Equivalent degree OR within 5 years of obtaining PhD degree depending on national context) and were aged between 25-45 years. All participants who gave informed consent to participate in online surveys and expert group discussions were included in the study. A sample size of a minimum of 100 respondents for phase I (online Google Form^LM^ survey) across at least ten countries in the world and a minimum of twenty respondents from phase I for phase II (online expert group discussion) were decided based on feasibility, time constraints, and their COVID-19 pandemic circumstances.

**Table 1:** Phase I Survey - Socio-demographic and addiction medicine training-related information (n=110)

| Variables | Mean (SD)/Frequency (%) |
| --- | --- |
| **Age (in years)** | 35.66 (4.97) |
| **Gender**<br>Male<br>Female | 68 (61.8 %)<br>42 (38.2 %) |
| **World-Health Organization (WHO) Regions**<br>Africa<br>Americas<br>Eastern Mediterranean<br>European<br>Southeast Asia<br>Western Pacific | 27 (24.5%)<br>14 (12.8%)<br>12 (10.9%)<br>16 (14.6%)<br>28 (25.4%)<br>13(11.8%) |
| **Type of professionals**<br>General/Addiction Psychiatrist<br>General/Addiction Physician<br>Clinical Psychologist<br>General/Psychiatrist Nursing<br>Psychiatric Social Worker<br>Doctoral (PhD) student/Post-Doctoral Fellow | 62 (56.5%)<br>30 (27.3%)<br>5 (4.5%)<br>4 (3.6%)<br>4 (3.6%)<br>5 (4.5%) |
| Years of clinical/research experience in Addiction Medicine | 5.78 (9.35) |
| Deficiency in existing addiction training programme/curriculum for ECAMPs | Yes – 95 (86.4%)<br>No – 15 (13.6%) |

### 2.3. Recruitment strategy

For the first phase of the study, potential participants were identified using the membership directory of organizations working in the field of addiction medicine {e.g., International Society of Addiction Medicine (ISAM), International Society of Substance Use Prevention and Treatment Professionals (ISSUP), World Psychiatric Association (WPA), Indian Psychiatry Society (IPS), and social media (LinkedIn) and research network (ResearchGate). We ensured that our sample included at least ten participants from each World Health Organization (WHO) region (African Region, Region of the Americas, South-East Asia Region, European Region, Eastern Mediterranean Region, and Western Pacific Region) in order to increase diversity and global representation. For the second phase of the study, all participants who took part in the first phase were randomized per WHO regions and were invited to participate in the in-depth interviews (within online expert group discussions) on the themes that emerged on training, research, and mentorship, using stratified random sampling strategy. A random sample of participants was engaged in 3 sessions of discussion, each comprising 8-11 respondents, for a duration of 2 hours, in April 2021. The participants were contacted in advance through email with enclosed information about the questionnaire, an expert group discussion guide, the rules of engagement in the discussion, the participant information sheet, and a consent form. Upon receiving written consent, a link for an online meeting was shared.

The online expert group discussions (Training, Research, and Mentorship) were facilitated by collaborator members from the ISAM NExT. The moderator (an ISAM NExT Member) guided the participants with questions and facilitated the discussion. The meetings were video recorded (with permission from the participants) and later transcribed for thematic analysis.

### 2.4. Ethics approval and consent

The online survey was conducted according to the principles of good scientific practice (15). Ethical approval for the study was sought and granted by the Institutional Ethics Committee (IEC) at the All India Institute of Medical Sciences (AIIMS), New Delhi, India (September 16, 2020, reference number IEC-888/04.09.2020). Participants provided a written online informed consent to participate in this study before filling out the self-administered survey, voluntary and anonymously.

### 2.5. Analysis

Data of phase I was analyzed using the Software Package for Social Sciences for Windows v. 24.0 (SPSS 24) (IBM Corp, Armonk, NY). Categorical variables were summarized as n (%), and continuous variables as means [standard deviation (SD)]. Focus group discussions data were analyzed using thematic analysis of the video recorded expert group discussions transcribed to the word document by the independent researchers (PK, KR). A core group of 13 collaborators from the ISAM NExT committee reviewed the data before publication.

## 3. Results

### 3.1. Phase I: Online Survey

Out of the 270 potential respondents approached, a total of 125 responses were received from across 56 countries and 6 WHO regions during six months of the data collection period (response rate 46.3%). Figure 1 shows the geographical distribution of the survey respondents (Phase 1) and the WHO regions. Fifteen responses were excluded as 10 respondents did not fulfill eligibility criteria and 5 responses were duplicates; therefore, data were analyzed for a total of 110 respondents for the phase I survey (Table 1). The mean age of respondents was 35.66 (SD=4.97) years. About half of the respondents were from the Southeast Asia and Africa region. Respondents had been working in the field of addiction medicine or psychiatry profession for an average of 5.78 years (ranging from 1-17 years).

**Figure 1.**
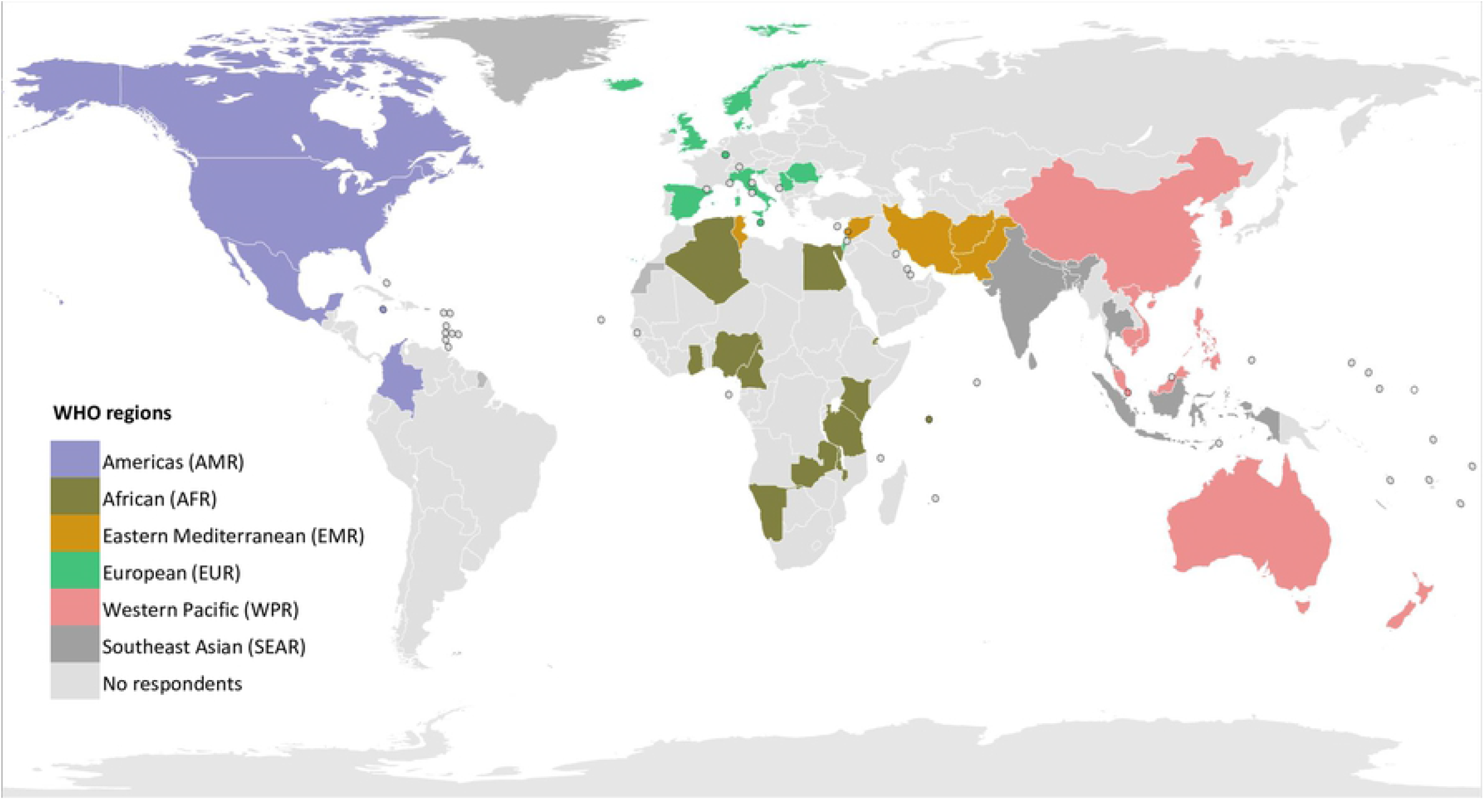
the geographical distribution of survey respondents (Phase I) and their affiliated WHO regions illustrated on a world map.

Out of these respondents, 56% were psychiatrists, with some exclusively practicing addiction psychiatry. Around 27% of respondents were addiction medicine physicians; psychologists (4.5%), nursing professionals (3.6%), social workers (3.6%), and post-doctoral fellows (4.5%) constituted the remaining professionals. Table 2 overviews the responses to the survey questions concerning key themes related to addiction medicine training, mentorship, and research shortages and needs.

**Table 2:** Phase I Survey – key themes for addiction medicine training needs.

|  |  |
| --- | --- |
| Key problems in lack of standardized training programmes | 1. Exposure to specialty clinics – 23 (24.2%)<br>2. Lack of trained faculty in subject – 18 (18.9%)<br>3. Less rotation/duration of posting in addiction medicine during postgraduate training – 18 (18.9%)<br>4. Lack of proper assessment of training – 14 (14.8%)<br>5. Lack of standard textbook – 14 (14.8%)<br>6. Less patient load – 8 (8.4%) |
| Lack of research opportunities for ECAMPs | Yes - 98 (89.1%)<br>No – 12 (10.9%) |
| Ways to improve research opportunities for ECAMPs | 1. Research project compulsory part of training – 28 (28.6%)<br>2. Workshops on how to conduct research – 26 (26.5%)<br>3. Workshops on how to get funding/grant for research – 20 (20.4%)<br>4. Funding (Intramural/Extramural) – 16 (16.3%)<br>5. Interdisciplinary team – 8 (8.2%) |
| Key recommendations for a successful mentorship programme | 1. Trained addiction professionals as mentors – 35 (31.8%)<br>2. Networking opportunities – 19 (17.3%)<br>3. Mentorship programme as an integral part of training – 18 (16.4%)<br>4. Institutional support – 17 (15.4%)<br>5. No recommendation – 21 (19.1%) |

### 3.2. Phase II: Online expert group discussions (n=28)

All participants of the phase I study were invited to participate in group discussions on training, mentorship, or research needs. A total of 110 participants were randomized based on the WHO regions (stratified) into groups of 37, 37, and 36. Subsequently, these groups were assigned one of the three themes (Training, Mentorship, and Research), and an email invitation along with questions on that theme was sent 10-14 days before the online expert group discussions. A total of 28 respondents participated in phase II (overall response rate 25.4 %). The response rate was 8/36 (22.2%) for the training theme, 8/37 (21.6%) for mentorship theme, and 12/37 (32.4%) for the research theme. The geographical distribution and the number of representatives in expert group discussions are depicted in figure 2. The thematic analysis results of the expert group discussions are as shown in Tables 3, 4, and 5.

**Figure 2.**
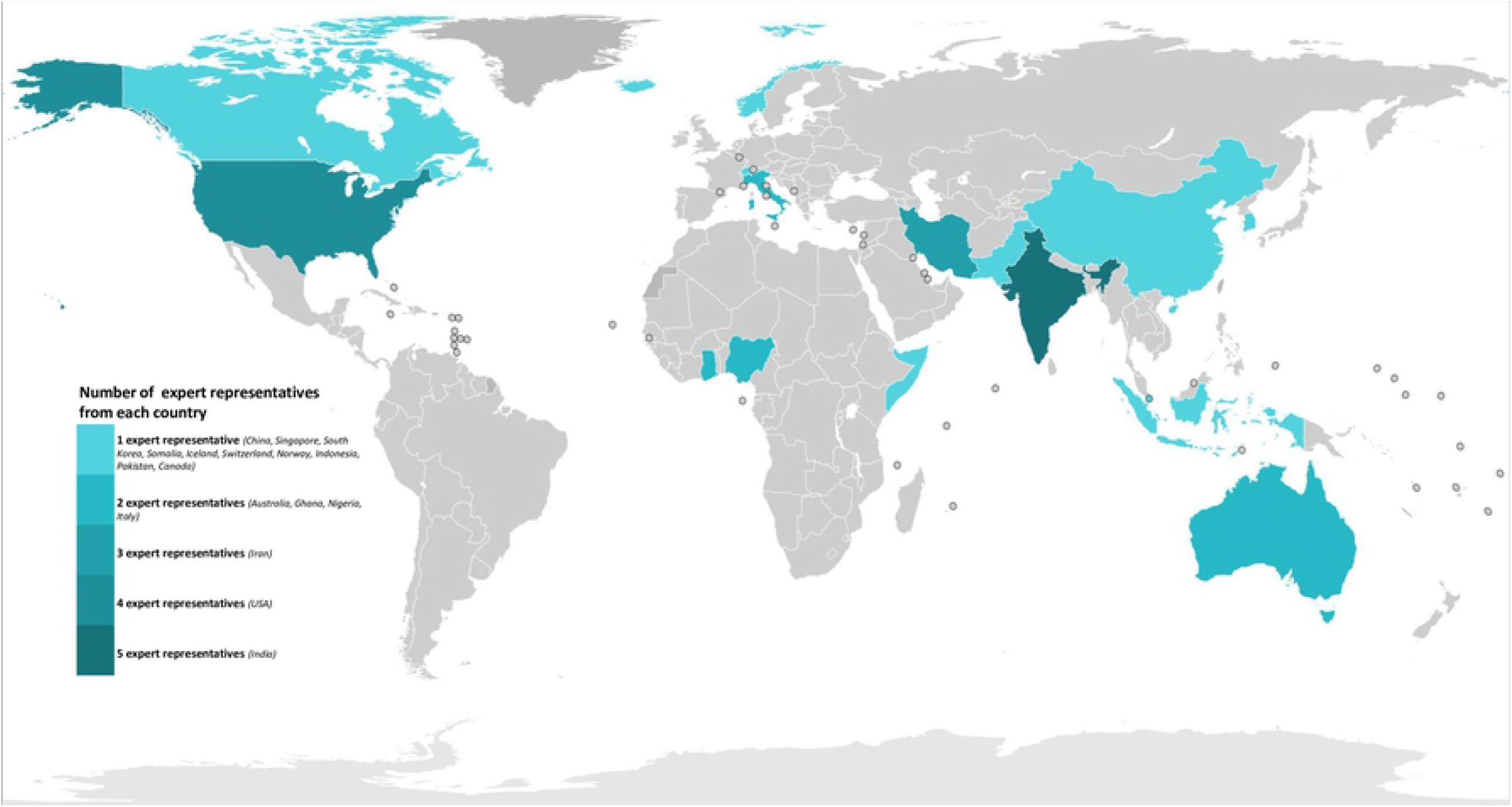
the number and geographical distribution of representatives who contributed to expert group discussions illustrated on a world map.

**Table 3A:** Expert group discussion on training (n=8)

| Part A: | Responses to the questions for advance distribution (frequency) |
| --- | --- |
| Countries (Participants) | Italy (2), India (2), Ghana (2), Indonesia (1), Iran (1) |
| Questions | Responses (n) |
| 1. Current phase of training in the field of Addiction Medicine/Psychiatry? | • Post-graduate/super-specialty Trainee Resident (3/8)<br>• Junior faculty member/Consultant (3/8)<br>• Nursing professional (2/8) |
| 2. Availability of standard training programme for Addiction Medicine/Psychiatry in medical school/institute/university? | • Yes (6/8)<br>• No (2/8) |
| 3. Duration of training in Addiction Medicine/Psychiatry in respondent's country | • Two years or less (1/6)<br>• Three years or less (4/6)<br>• More than three years (1/6) |
| 4. How satisfied respondent is with the training programme in addiction medicine/psychiatry | • Very unsatisfied (1/6)<br>• Neutral (1/6)<br>• Satisfied (3/6)<br>• Very satisfied (1/6) |
| 5. Main strengths of the training programme | • Exposure to the management of different substances/specialty clinics (4/6)<br>• Availability of eminent addiction professionals as mentors (2/6) |
| 6. Main limitations of the training programme | • Lack of addiction specialists as trainers (4/6)<br>• Lack of specialty clinics and exposure to multiple substance use problems (1/6)<br>• Difficulties in obtaining needed materials for training (1/6) |
| 7. Number of patients consulted under supervision/year during training? | • Less than 10 patients/year (1/6)<br>• 20-50 patients/year (1/6)<br>• More than 50 patients/year (4/6) |
| 8. Exposure/posting/rotation to Addiction Medicine/Psychiatry during medicine or psychiatry training? | • Yes (8/8) |
| 9. Duration of posting/rotation in Addiction Medicine/Psychiatry | • One month or less (2/8)<br>• 1-3 Months (3/8)<br>• 3-6 months (3/8) |
| 10. Exposure/clinical experience in the following subspecialties/special treatment facilities of Addiction Medicine/Psychiatry | • Dual diagnosis clinic, Opioid agonist treatment, Behavioural addiction clinic (7/8)<br>• Consultation liaison (6/8)<br>• Tobacco cessation clinic, Adolescent clinic (5/8)<br>• Community addiction treatment clinic, Injecting drug users clinic (4/8)<br>• Pain clinic, New psychoactive substances (NPS) clinic (1/8) |

**Table 3B:**
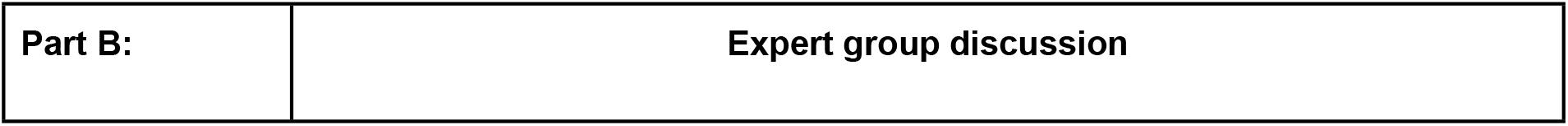
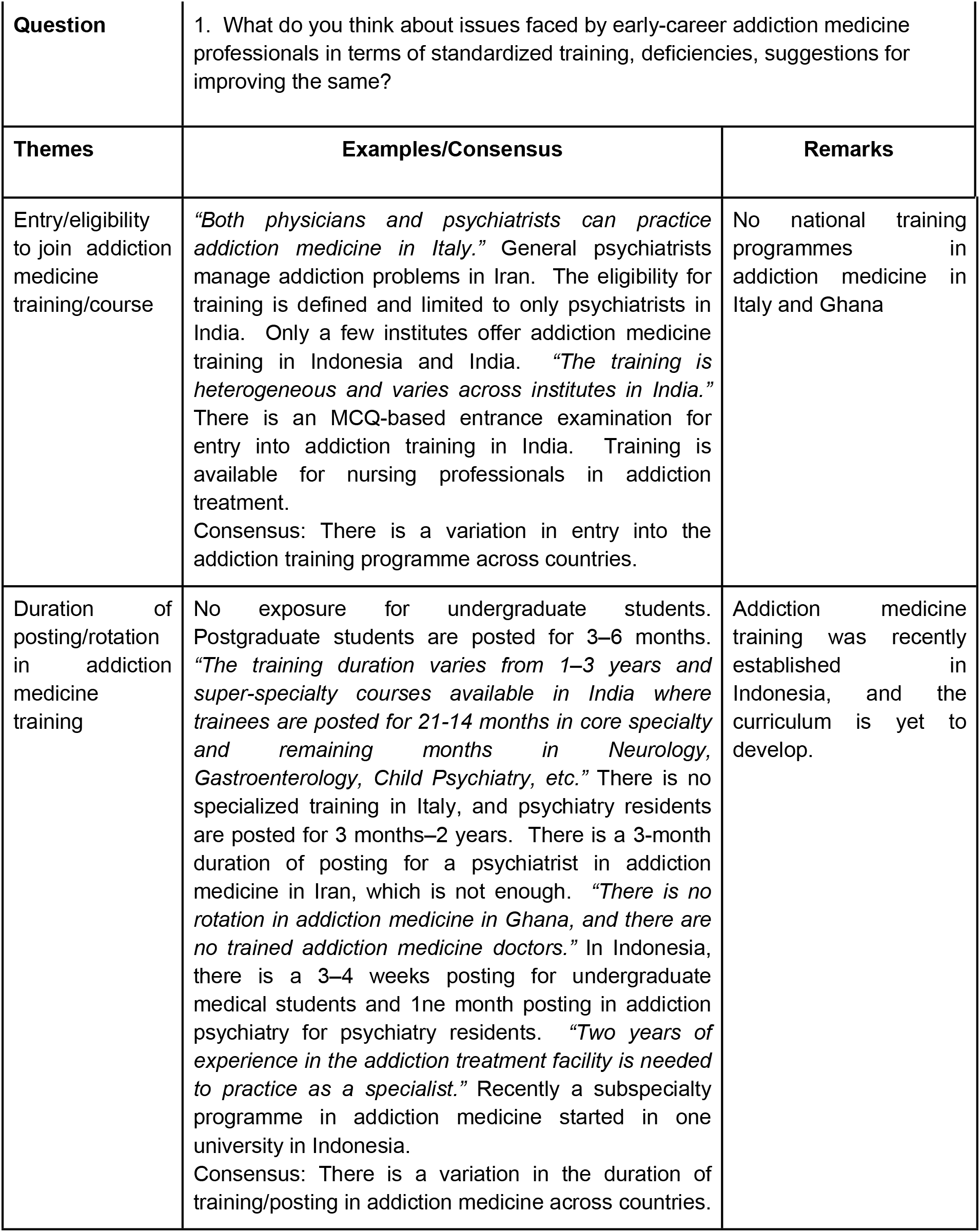
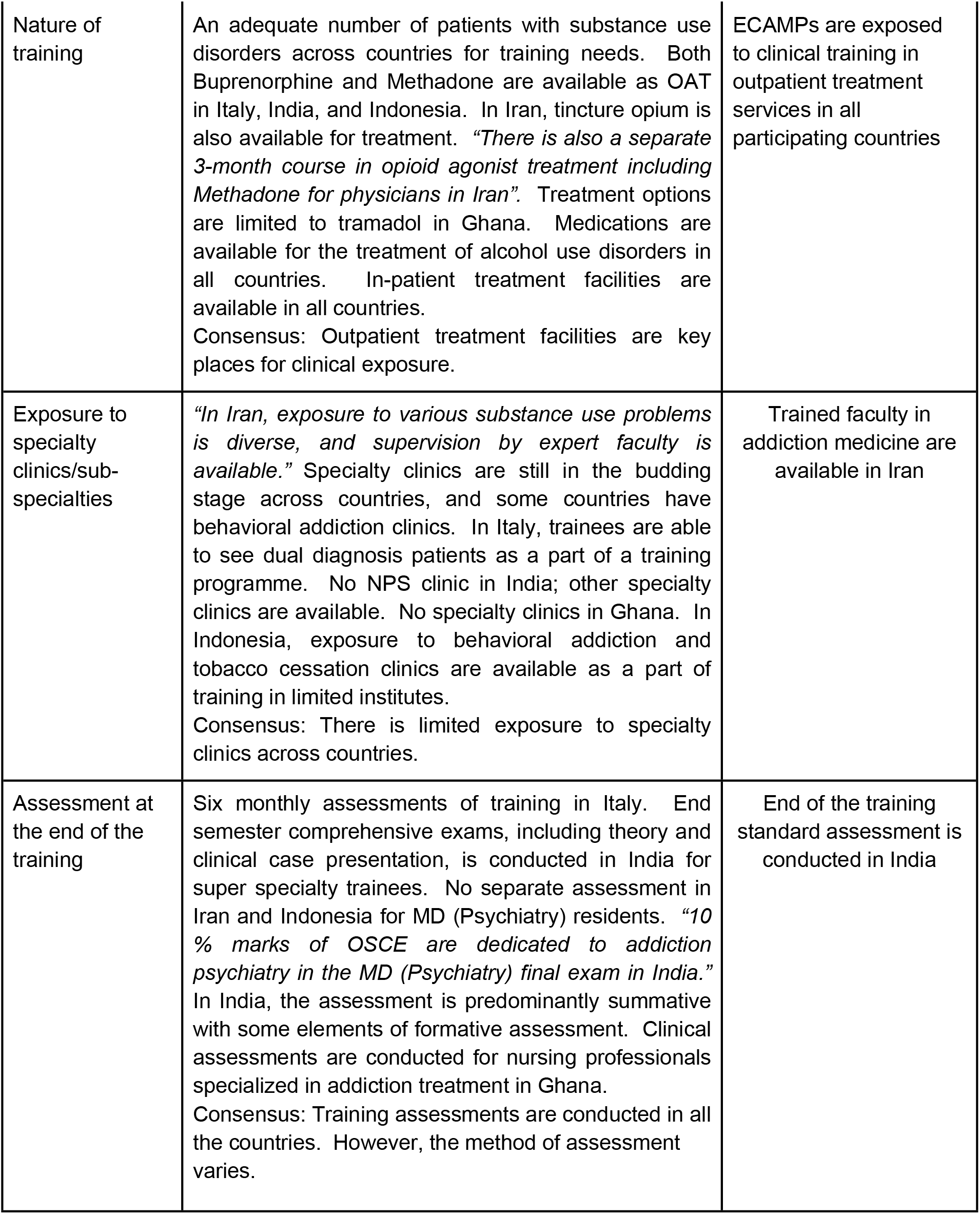
Thematic analysis of expert group discussion (Training)

|  |  |
| --- | --- |
| **Part B:** | **Expert group discussion** |

|  |  |  |
| --- | --- | --- |
| **Question** | 1. What do you think about issues faced by early-career addiction medicine professionals in terms of standardized training, deficiencies, suggestions for improving the same? |  |
| **Themes** | **Examples/Consensus** | **Remarks** |
| Entry/eligibility to join addiction medicine training/course | *“Both physicians and psychiatrists can practice addiction medicine in Italy.”* General psychiatrists manage addiction problems in Iran. The eligibility for training is defined and limited to only psychiatrists in India. Only a few institutes offer addiction medicine training in Indonesia and India. *“The training is heterogeneous and varies across institutes in India.”* There is an MCQ-based entrance examination for entry into addiction training in India. Training is available for nursing professionals in addiction treatment.<br>Consensus: There is a variation in entry into the addiction training programme across countries. | No national training programmes in addiction medicine in Italy and Ghana |
| Duration of posting/rotation in addiction medicine training | No exposure for undergraduate students. Postgraduate students are posted for 3–6 months. *“The training duration varies from 1–3 years and super-specialty courses available in India where trainees are posted for 21-14 months in core specialty and remaining months in Neurology, Gastroenterology, Child Psychiatry, etc.”* There is no specialized training in Italy, and psychiatry residents are posted for 3 months–2 years. There is a 3-month duration of posting for a psychiatrist in addiction medicine in Iran, which is not enough. *“There is no rotation in addiction medicine in Ghana, and there are no trained addiction medicine doctors.”* In Indonesia, there is a 3–4 weeks posting for undergraduate medical students and 1ne month posting in addiction psychiatry for psychiatry residents. *“Two years of experience in the addiction treatment facility is needed to practice as a specialist.”* Recently a subspecialty programme in addiction medicine started in one university in Indonesia.<br>Consensus: There is a variation in the duration of training/posting in addiction medicine across countries. | Addiction medicine training was recently established in Indonesia, and the curriculum is yet to develop. |
| Nature of training | An adequate number of patients with substance use disorders across countries for training needs. Both Buprenorphine and Methadone are available as OAT in Italy, India, and Indonesia. In Iran, tincture opium is also available for treatment. *"There is also a separate 3-month course in opioid agonist treatment including Methadone for physicians in Iran"*. Treatment options are limited to tramadol in Ghana. Medications are available for the treatment of alcohol use disorders in all countries. In-patient treatment facilities are available in all countries.<br>Consensus: Outpatient treatment facilities are key places for clinical exposure. | ECAMPs are exposed to clinical training in outpatient treatment services in all participating countries |
| Exposure to specialty clinics/sub-specialties | *"In Iran, exposure to various substance use problems is diverse, and supervision by expert faculty is available."* Specialty clinics are still in the budding stage across countries, and some countries have behavioral addiction clinics. In Italy, trainees are able to see dual diagnosis patients as a part of a training programme. No NPS clinic in India; other specialty clinics are available. No specialty clinics in Ghana. In Indonesia, exposure to behavioral addiction and tobacco cessation clinics are available as a part of training in limited institutes.<br>Consensus: There is limited exposure to specialty clinics across countries. | Trained faculty in addiction medicine are available in Iran |
| Assessment at the end of the training | Six monthly assessments of training in Italy. End semester comprehensive exams, including theory and clinical case presentation, is conducted in India for super specialty trainees. No separate assessment in Iran and Indonesia for MD (Psychiatry) residents. *"10 % marks of OSCE are dedicated to addiction psychiatry in the MD (Psychiatry) final exam in India."* In India, the assessment is predominantly summative with some elements of formative assessment. Clinical assessments are conducted for nursing professionals specialized in addiction treatment in Ghana.<br>Consensus: Training assessments are conducted in all the countries. However, the method of assessment varies. | End of the training standard assessment is conducted in India |

**Table 4A:** Expert group discussion on mentorship (n=8)

| Part A: | Responses to the questions sent for advance distribution |
| --- | --- |

| Countries (Participants)/ Questions | Canada (1), Iran (1), India (1), Nigeria (1) Singapore (1), Somalia (1), USA (2) |
| --- | --- |
| Questions | Responses (n) |
| 1. How would you describe your current phase of career in the field of addiction medicine? | ● Post-graduate/super-specialty Trainee Resident (3/8)<br>● Junior faculty member (2/8)<br>● Doctoral/Fellowship Student (1/8)<br>● Post-doctoral Student/Fellow (1/8)<br>● Associate Physician (1/8) |
| 2. Do you have a formal mentorship programme in your medical school/institute/university? | ● Yes (3/8)<br>● No (5/8) |
| 3. Do you consider your current supervisor/thesis guide/principal investigator as your mentor? | ● Yes (5/8)<br>● No (2/8)<br>● No, but I have a mentor from outside my medical school/institute/university (long-distance mentor) (1/8) |
| 4. If a mentorship programme was available, then how satisfied were you with the mentorship programme? | ● Unsatisfied (1/3)<br>● Neutral (1/3)<br>● Satisfied (1/3) |
| 5. Can you describe two reasons for effectiveness/non-effectiveness (if any) for the available mentorship programme? | Effectiveness<br>● Similar experiences, life goals, and professional aims to mine, an ability to relate (2/3)<br>● Genuine concern for my wellbeing and success (1/3)<br>Non-effectiveness<br>● Limited time and busy schedule, and lack of protected time for mentoring and research (2/3)<br>● Mentors possessed biased views against non-psychiatrist's ability to practice in the field of addiction (1/3) |
| 6. Ideally, how many mentors should a mentee have? | • Single mentor (5/8)<br>• Multiple mentors (2/8)<br>• Multiple mentors with a single senior mentor in charge (1/8) |
| 7. According to your experience, which phase of a mentorship programme* is the most important to you?<br>* Zachary's The Mentor's Guide, 2000 (15) | • Initiation or Preparation (1/8)<br>• Cultivation or Negotiation & Enabling or Protégé (5/8)<br>• Separation or Closing or Break up (1/8)<br>• Redefinition or Lasting friendship (1/8) |
| 8. Do you have an office for delivering needed guidance on career development/training/post-doctoral while training at your medical school/institute/university? | • Yes (1/8)<br>• No (7/8) |
| 9. If the ideal mentorship programme is available, then what sort of guidance and assistance would you need from a mentor as a mentee (in order of preference)? | • Long-term career planning (3/8)<br>• Research design (1/8)<br>• Networking nationally and internationally (1/8)<br>• Balancing personal & Professional demands (1/8)<br>• Developing a research portfolio (1/8)<br>• Addressing burnout during training (1/8) |
| 10. By the end of an effective mentorship programme, what kind of abilities should a mentee obtain? | • Ability to conduct research ethically independently and responsibly in the given area (4/8)<br>• Ability to achieve career progression financial independence through satisfactory job opportunities in the given area (3/8)<br>• Ability to become a mentor and run an effective mentorship programme in the given area (1/8) |

**Table 4B:** Thematic analysis of expert group discussion (Mentorship)

| Part B: | Expert group discussion |  |
| --- | --- | --- |
| Questions | 1. In Your university, in your city, in your academic field, do you have a mentorship programme in the field of addiction medicine or general psychiatry? If so, how satisfying is the programme? |  |
| Themes | Examples/Consensus | Remarks |
| Availability of mentorship programme | *"There is no formal mentorship programme in some institutes. But anyone can identify someone with similar research interests as a mentor. There is a mentorship programme for Buprenorphine training to get a special waiver to prescribe Buprenorphine in USA (Called as PCSS-O), but it is not associated with any university, and it is not that satisfying."*<br>Consensus: The mentorship programme is not available widely in developed countries and non-existent in developing countries | Mentorship programmes for early-career addiction medicine professionals are not available in most countries |
| Nature of mentorship programme | *"In Canada, there is mentorship programme only for current fellows in addiction medicine, for the duration they are in the programme to provide them primarily with career advice."*<br>Consensus: There are no formal mentorship programmes for addiction medicine in most institutes, and among the available programmes, only a few are satisfactory. |  |
|  | 2. Can you describe reasons for effectiveness and non-effectiveness for mentorship programmes? |  |
| Effectiveness/non-effectiveness of the mentorship programme | *"Mismatch about expectations is a very critical thing. There should be flexibility in the programme for people or prospective mentees to be specific in what they are looking for and choose mentors based on their expectations. The relationship between mentor and mentee is another important reason for failure or success of a programme."*<br>Consensus: The programme should be interdisciplinary, and mentees should be able to choose their mentor based on their needs or interests; The mentor should have adequate emotional intelligence, should be available and accessible, and be able to relate to the mentees, and there should be SMART goals. The programme should not be short-term but should be continuous or long-term. |  |
|  | 3. What are your suggestions for successful mentorship programmes, especially for early-career addiction medicine professionals? |  |
| Appropriate time and duration for the mentorship programme | *"Exposing medical students early in their career, preferably at the undergraduate level even before they decide to go into psychiatry or surgery specialties, etc. Identifying those who are passionate about addiction. For graduates or those in private practice, etc., ISAM or MSR can create networks. There is a need for continuity instead of being short-lived. Information about mentorship programmes should be given to trainees during the last year of training."* |  |
| Career-based mentorship programme | *"We need to have early career mentoring programmes. Successful mentoring programmes should have time-bound SMART goals. Also, some kind of career-based mentoring on how to maintain a balance between personal and professional aspects of life. We should have some wellness programmes on how to balance lifestyle and work in addiction."*<br>Consensus: For a successful mentorship programme, it should be introduced at the medicine level (undergraduate level), residency as well as early-career level. There should be flexibility keeping mentees in the center of everything, and mentees should be able to choose their mentors. There should be SMART goals. The programme should not be short-lived but should be continuous and comprehensive. ISAM can help in international collaboration for mentorship. Also, there should be a career-based mentoring programme to discuss work-life balance issues. | Continuous mentorship programme as per the stage of career is needed for its effectiveness in shaping the career of ECAMPs |

**Table 5:**
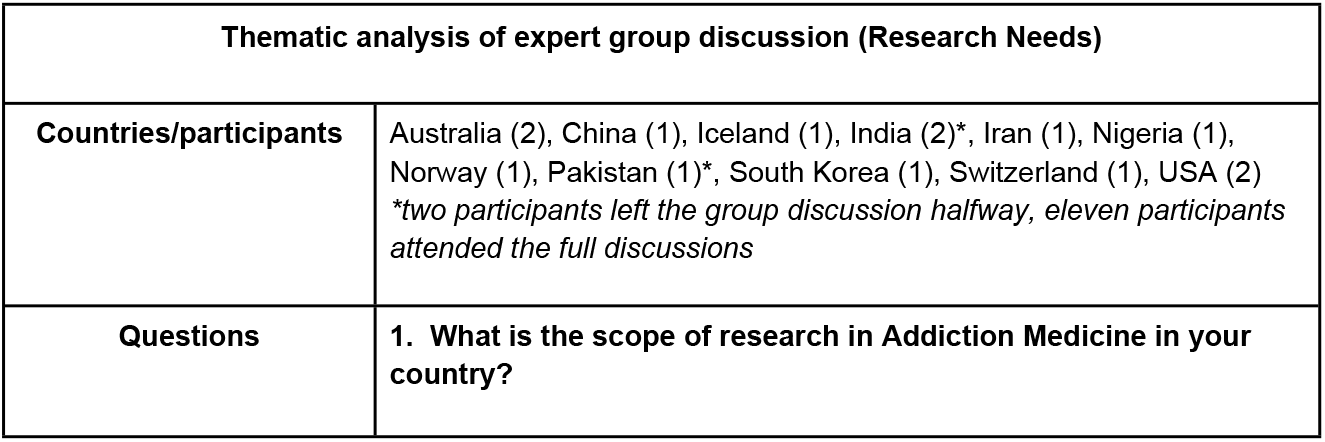
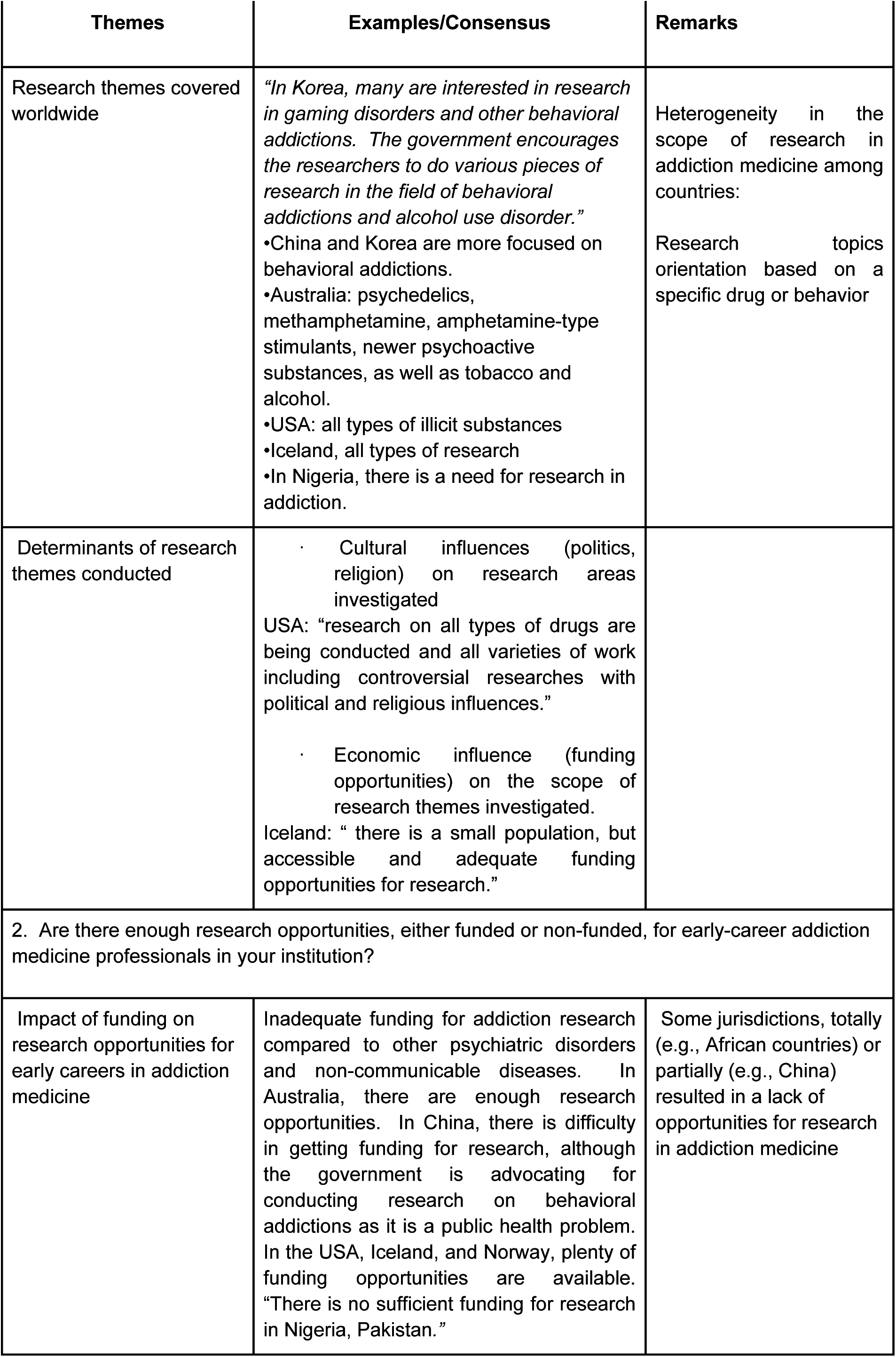
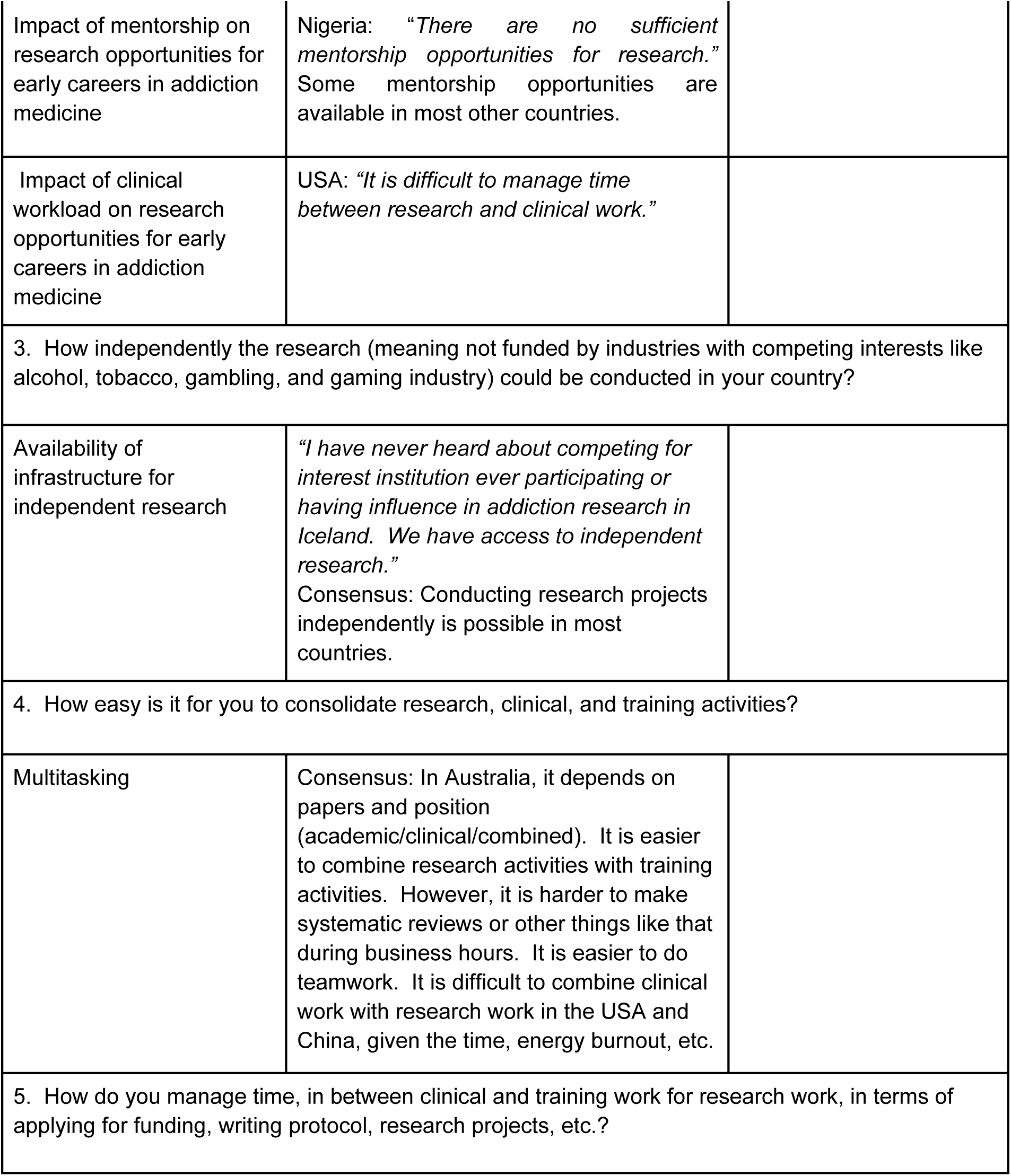
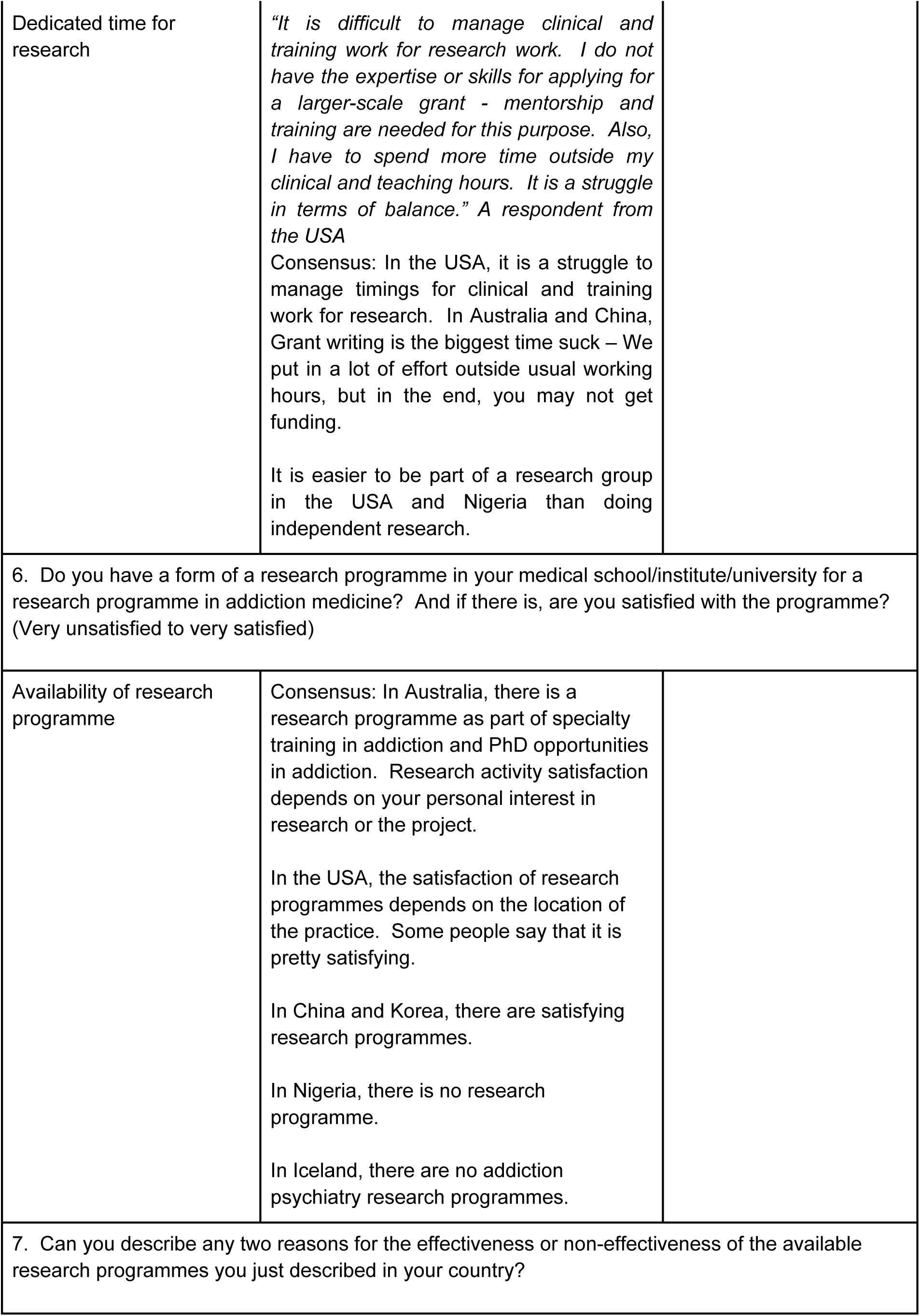
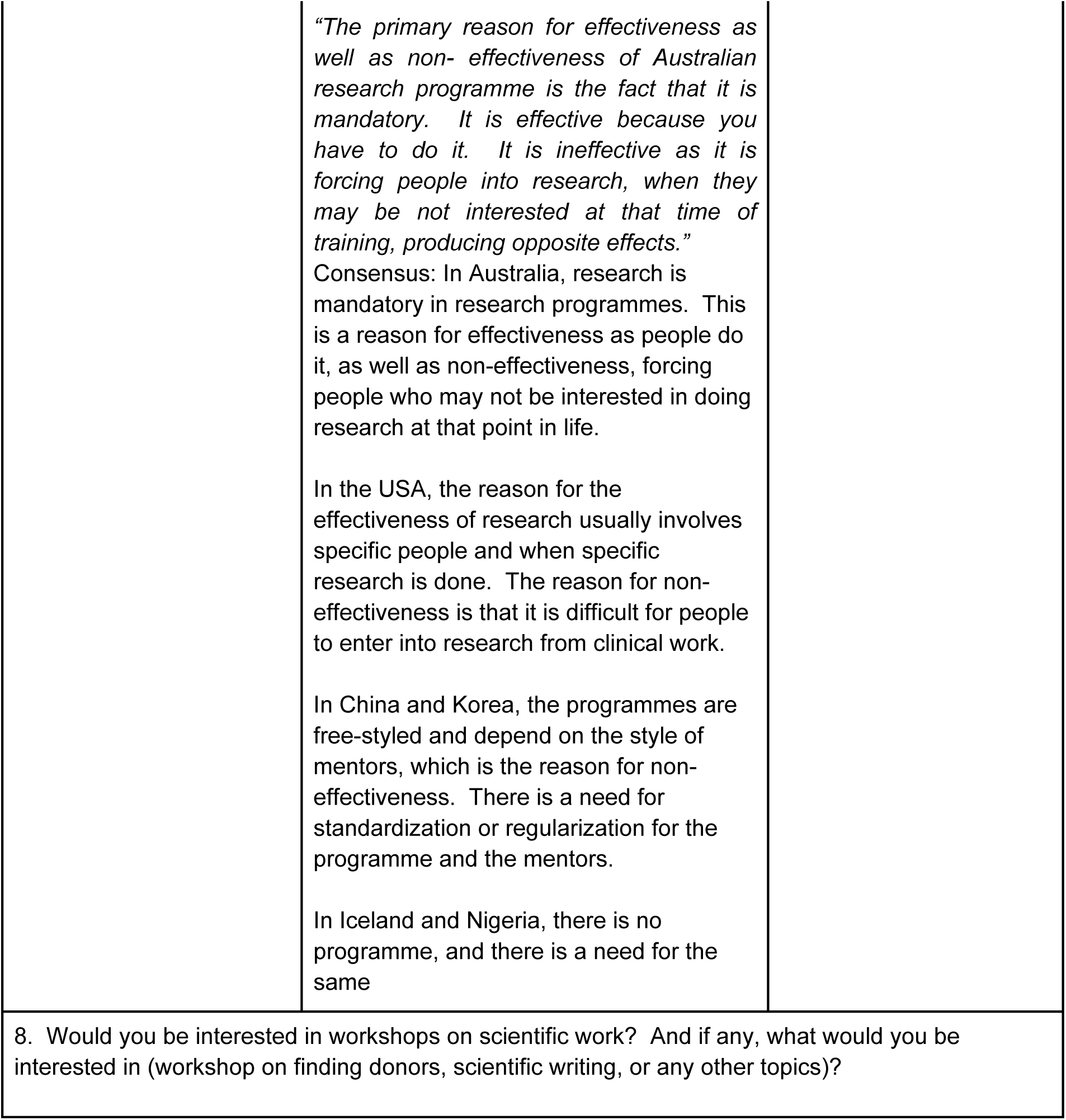
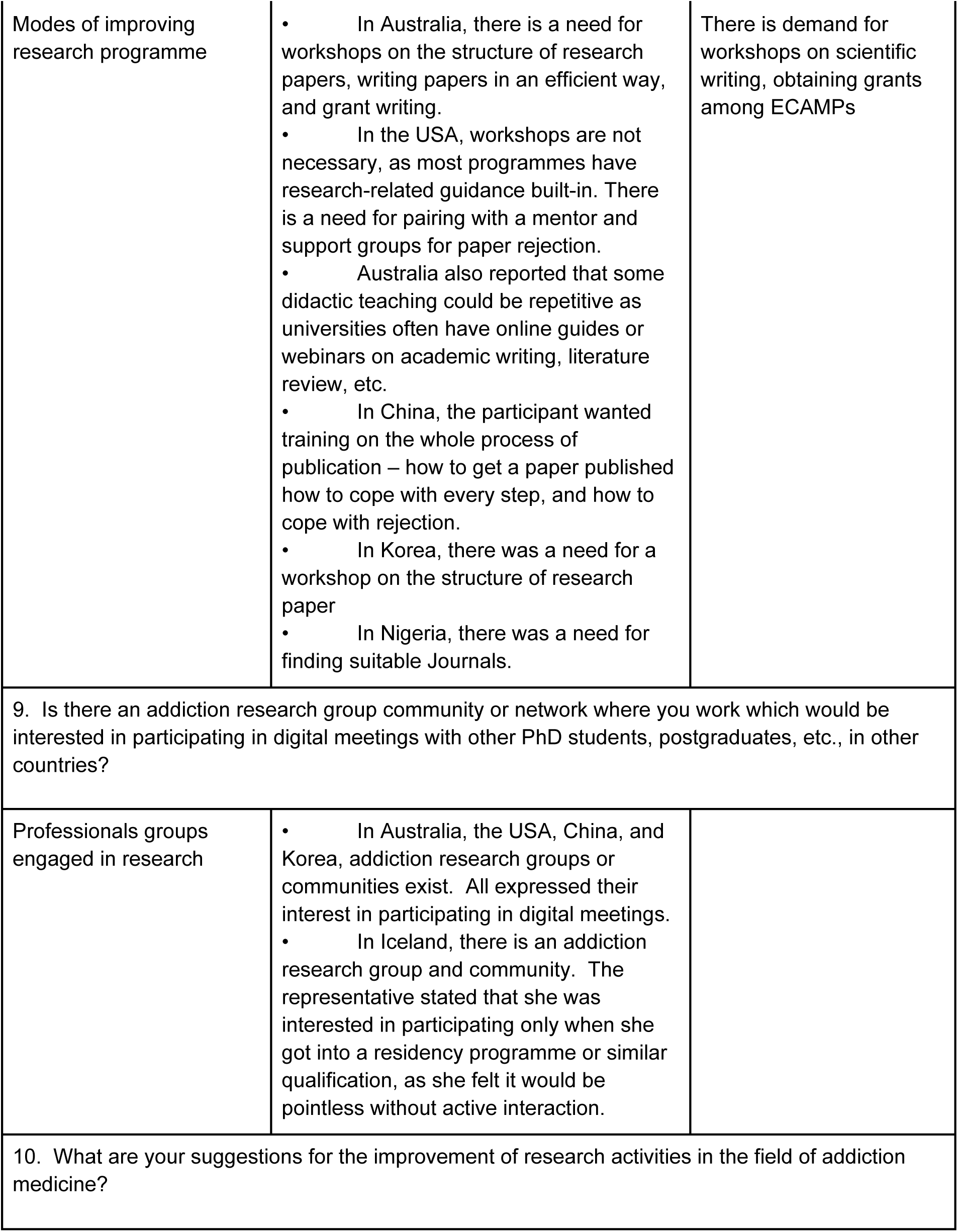
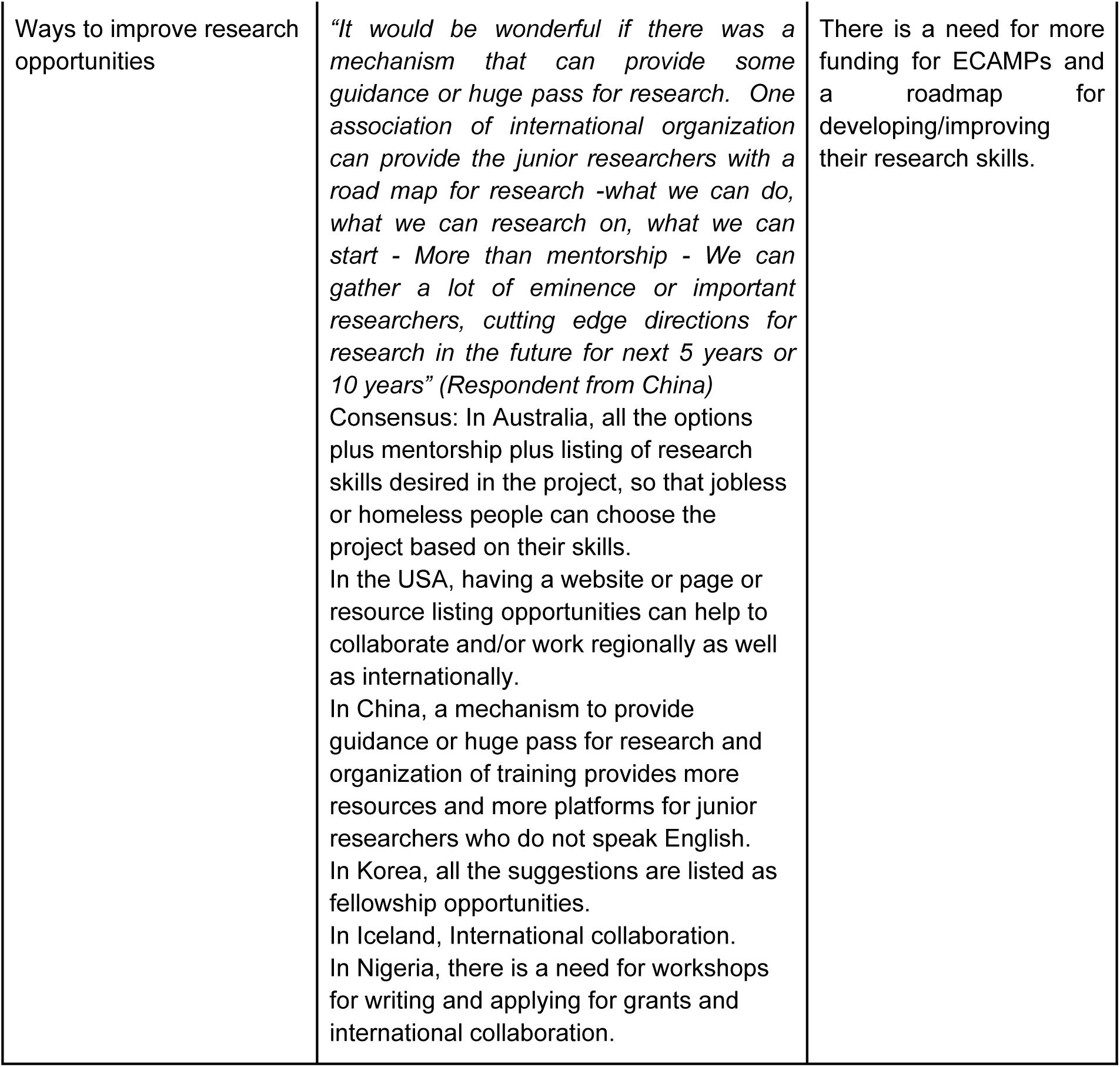
Expert group discussion on research needs (n=12)

## 4. Discussion

This study was one of the most extensive surveys conducted among early-career addiction medicine professionals assessing the need and scope for standardized training, mentorship programmes, and research opportunities — the online survey methodology allowed for a broad representation of participants from 56 countries.

There is wide variability in entry requirements for addiction medicine training globally. In countries such as the USA and the UK, both family physicians and psychiatrists can practice as addiction specialists. In others, such as in Italy, both physicians with a specialty in internal medicine or other medical subspecialties together with psychiatrists and pharmacologists can treat patients with addiction. In most other countries, only psychiatrists can train in addiction medicine as a specialty. Developing countries allow nursing practitioners and social workers to pursue addiction medicine training due to a shortage of specialty physicians.

Training and exposure to addiction medicine also differ considerably across different countries. With respect to undergraduate training, exposure to addiction medicine as a specialty is minimal (about one week in the USA, 7 hours in the UK) or absent in most countries (6, 16–20). The biopsychosocial model of addiction is taught as a part of the theory in undergraduate medical school (21). Addiction medicine is an integral part of psychiatry and family medicine residencies in some countries like the USA, India, and Iran with respect to postgraduate training. Indonesia and India offer additional certificate courses after postgraduate training. There is considerable variation in curriculum and duration of the training placement of psychiatry residents in addiction psychiatry across countries.

In the USA & Australia, addiction psychiatry is a separate specialty as a postgraduate programme. In India and Indonesia, addiction psychiatry specialty is in the early developing stage (18,20,22). Lack of a structured curriculum is an important issue highlighted by the present survey participants. Content of the addiction medicine curriculum varies due to the nature of local substance use, availability of specialty clinics, opioid agonist treatments and other pharmacotherapy options available in different countries, and availability of trained faculty members for teaching. A structured curriculum can improve the knowledge of addiction medicine among internal medicine residents and hence need to be developed updated in different countries in order to improve the delivery of quality addiction treatment services (23). The quality of training is an issue for both internal medicine residents in the USA and also among psychiatry trainees across European countries as per a recent survey, which makes the call for a structured curriculum ever more important and urgent (24,25).

A survey in China revealed that doctors involved in drug treatment are not well prepared or experienced and have negative attitudes toward substance use disorders and afflicted patients (26). The low number and level of professional addiction experts are the potential outcomes of inadequate addiction medicine training for medical students and residents in the USA, which is highlighted previously (9). From a trainee point of view, there is a demand for standardized training as emphasized in past reviews and found in the present study (27,28). The evaluation of standardized, structured short term training is also found to be an effective tool for addiction medicine training (29, 30) . The next generation of addiction treatment providers needs to be trained adequately to deal with emerging substance use problems across the globe.

There is a standard exit exam after completion of addiction medicine training in some countries like the USA, Australia, and India. There are regular mid-term assessments (6 monthly or yearly) that are also conducted in countries like Italy, India, Iran, and Ghana for the trainees. In this regard, efforts by the ISAM to successfully conduct International Certification in Addiction Medicine exams for global trainees for the past 10 years need to be acknowledged (8).

The availability of mentorship programmes and needs were assessed in the present study. We found there are limited mentorship programmes available for early-career addiction medicine professionals. Such programmes are limited to developed countries like the USA & Australia. The mentorship programme is non-existent in most African and Asian countries like Ghana, Nigeria, China, India, Indonesia etc. Most participants recognized their training programme supervisor, thesis guide as a mentor. A single mentor was desired by most, although some participants expressed the need for multiple mentors depending upon the need in particular areas of interest and the stage of their career. The barriers identified for quality mentorship programmes were lack of time, funding, and trained faculty members (31). Most participants favored a continuous mentorship programme in different stages of their careers and were not limited to only the training duration. Mentorship programmes are vital for the development of the career of ECAMPs, and there is a need to facilitate mentorship programmes across nations. There are limited research studies on understanding the challenges faced with mentorship problems for ECAMPs. Among the available programmes, The Learning for Early Careers in Addiction & Diversity (LEAD) Programme, funded by the National Institute of Drug Abuse, uses a team mentoring approach. Each LEAD Programme scholar works with a Clinical Trial Network (CTN) primary mentor while also receiving guidance from a UCSF University of California San Francisco (UCCSF) mentor and a nationally regarded diversity advisor (32). Other similar programmes are run by addiction medicine societies like the American Academy of Addiction Psychiatry (AAAP) & ISAM (International Society of Addiction Medicine) (33). There is a tremendous need to develop the culture of mentorship for strengthening academic medical centers engaged in addiction medicine training. Innovative methods like co-training with general physicians can facilitate mentorship programmes in such centres. The mentoring need is now even greater with the expansion of addiction medicine as a specialty and many young professionals joining their respective training programmes (11, 34, 35).

Most of the study participants reported there are limited research opportunities for ECAMPs. There are many challenges like clinical workload, funding, few suitable research mentors, obtaining research grants, and publishing the research. The challenges are existent even in developed countries like the USA & Australia. The research capacity has to be more developed during the training programme and is effective when mandatory for the completion of training. There is an unmet demand for grant writing, workshops for conducting research, and writing papers among ECAMPs. The research grants available from NIDA are mostly limited to USA Residents/Citizens (10). There are limited opportunities from addiction medicine societies. The United Nations Office on Drugs & Crime (UNODC), with support from the Drug Abuse Prevention Center (DAPC), started offering grants for early career researchers for projects related to prevention and promotion activities recently (36).

Combining clinical training and research would be a step ahead in improving addiction medicine training programmes and creating research opportunities for ECAMPs (37). Developing research capacity among ECAMPs from low and lower-middle-income countries by conducting workshops with the support of facilitators from high-income countries can be a solution for the problem. Other allied addiction medicine professionals can also be engaged in such training programmes to develop workforce and build more capacity (35, 38, 39). The main challenges encountered for conducting research by ECAMPs in the European survey (n=258) were lack of time as a large proportion of participants (87.2%) reported conducting research after regular working hours or partly during and after working hours. Only one-tenth ever received a grant for their work. Lack of funding is an important hurdle in conducting research in spite of ECAMPs being motivated to conduct the research(40). Global societies and institutes working in the field of addiction medicine need to provide adequate research opportunities as there is a risk of early-career addiction medicine professionals falling prey to predatory publishing and industry-sponsored research in the early stage of their career, which may bias their subsequent research projects (41–43).

The results from the present study suggest that there is variation in eligibility, the content of the curriculum, and assessments for addiction training across the globe. It is essential to develop a standard curriculum and training content that is competency-based, culturally sensitive, and can include local jurisdictional norms with substance use disorders. Flexibility is needed in the curriculum to account for the possibility of various medical professionals starting addiction medicine as a career. The study findings emphasized the need for mentorship programmes and more research opportunities for ECAMPs as a vital component of addiction medicine training.

A major strength of the present study is the perspective from more than 50 countries and covering all 6 WHO regions. We used a robust methodology with an online two-phase survey with systematic randomization for the second phase. The second phase, i.e., the qualitative part of the study using expert group discussions, adds perspective on attitudes and opinions of survey participants and hence adds more meaning and depth to the data collected using the online survey. Limitations of our study include self-reported data and a relatively small sample size. The study was conducted during the COVID-19 pandemic, where there was disruption of training programmes and shift to online teaching, which may have influenced some of the findings in the study. The generalizability of the data is another limitation, as only participants who were members of professional societies and available on social media were approached. Future studies should address these limitations using randomized control trials for studying models of training, innovative techniques of training, and longitudinal study design to study mentorship needs in long-term career growth. The study findings emphasize the need for standardized training programmes, improving research opportunities and collaboration, and effective mentorship programmes for the next generation of addiction medicine professionals.

## Data Availability

All relevant data are within the manuscript and its Supporting Information files.

## Authors’ Contributions

RB & PK drafted the protocol. RB, MK, PR, and VLN recruited the phase I survey participants. PR sent the invitations and arranged online expert group discussion meetings. SA, MF, KM, PR developed questions and animated the discussions for expert group discussion research theme. RB, SA, MF, KM, PR, HMA, JLB, ST, MK, LO, VLN were collaborators for expert group discussions and facilitated the sessions. PK was note taker for expert group discussion sessions, transcribed and analyzed research & mentorship expert group discussions. KR validated the expert group discussions data. PR illustrated the figures and maps. ISAM NExT Consortia members participated in both phases of the survey. MNP, HE & AB reviewed the manuscript and mentored the group. All authors contributed to the revision and final edits of the manuscript.

## Acknowledgments

We would like to thank Dr Shalini Arunogiri, Training Officer, ISAM; and Dr Cor De Jong Co-chair, Training Committee, ISAM; Dr Susana Galea Singer, Education Officer, ISAM; Dr Nady el-Guebaly, Chief Examiner, ISAM for their guidance and Ms Marilyn Dorozio, ISAM NExT office for technical support in conducting online meetings.

